# Analysis of PTRHD1 common and rare variants in European patients with Parkinson’s disease

**DOI:** 10.1101/2020.12.11.20243402

**Authors:** Yuri L. Sosero, Sara Bandres-Ciga, Ziv Gan-Or, Lynne Krohn, on behalf of the IPDGC

**Affiliations:** Department of Human Genetics, McGill University, Montréal, QC, H3A 1A1, Canada; Montreal Neurological Institute, McGill University, Montréal, QC, H3A 1A1, Canada; Molecular Genetics Section, Laboratory of Neurogenetics, National Institute on Aging, National Institutes of Health, Bethesda, MD 20892, USA; Department of Neurology and Neurosurgery, McGill University, Montréal, QC, H3A 1A1, Canada

**Keywords:** PTRHD1, Parkinson’s disease, familial parkinsonism, burden test, SKATO, GWAS

## Abstract

Three family studies identified three different variants in the peptidyl-tRNA hydrolase domain containing 1 gene (*PTRHD1*) in patients affected by syndromic parkinsonism. In the current study, our objective was to investigate whether *PTRHD1* variants are associated with Parkinson’s disease (PD) risk and age at onset (AAO). To evaluate the association between *PTRHD1* and PD risk, we analyzed whole genome sequencing (WGS) data of 1,647 PD cases and 1,050 healthy controls, as well as genome-wide imputed genotyping data on 14,671 PD cases and 17,667 controls, all of European ancestry. Furthermore, we examined the association of *PTRHD1* with PD risk and AAO using summary statistics data from the most recent PD genome-wide association study (GWAS) meta-analyses. Our results show no association between *PTRHD1* and PD risk or AAO. We conclude that *PTRHD1* does not play a major role in PD in the European population. Further large-scale studies including subjects with different ancestry and family trios might further clarify the relationship of this gene with PD and atypical parkinsonism.

## 1. Introduction

Parkinson’s disease is a complex neurodegenerative disorder resulting from a variety of genetic and non-genetic risk factors. Three family studies nominated peptidyl-tRNA hydrolase domain containing 1 gene (*PTRHD1*) as a possible disease-causing gene in atypical parkinsonism (Jaberi et al., 2016; Khodadadi et al., 2017; Kuipers et al., 2018). In these studies, two conducted in Iranian families (Jaberi et al., 2016; Khodadadi et al., 2017) and one in a specific sub-Saharan African community (Kuipers et al., 2018), the carriers showed some level of early onset cognitive impairment, and later developed symptoms of parkinsonism. In particular, homozygous *PTRHD1* c.157C>T (p.His53Tyr) variants were detected in two Iranian siblings with intellectual disability and motor abnormalities. At their second/third decade of age, the siblings presented with muscle stiffness, anxiety, resting and postural tremor, along with hypersexuality and hypersomnia (Khodadadi et al., 2017). Similar symptoms, including intellectual disability/attention deficits and, successively, gait disturbance, bradykinesia, tremor and falls, have also been reported in two male siblings from another Iranian family, homozygous carriers of a *PTRHD1* c.155G>A (p.Cys52Tyr) variant (Jaberi et al., 2016). Likewise, the study on the Sub-Saharan Xhosa-speaking community showed that, within the same family, three homozygous carriers of a *PTRHD1* deletion (c.169_196del, p.Ala57Argfs*26) had mental disability and symptoms of parkinsonism, with a wider clinical variability compared to the two previous studies (one member showed just tremor, one just bradykinesia and hypokinesia, one both) (Kuipers et al., 2018).

Despite these findings, no large-scale study has been performed before in the European population to investigate the relationship between *PTRHD1* and PD. To test this potential association, we examined the *PTRHD1* region in Europeans using whole genome sequencing (WGS) data from the Accelerating Medicines Partnership - Parkinson’s disease initiative (AMP-PD; www.amp-pd.org) and large-scale genome-wide imputed genotyping data from the International Parkinson’s Disease Genomics Consortium (IPDGC).

## 2. Methods

### 2.1 Population

To investigate the association of *PTHRD1* common and rare variants with PD, we analyzed AMP-PD WGS data including 1,647 PD cases and 1,050 healthy controls of European descent from three different cohorts: Parkinson’s Disease Biomarker Program (PDBP; https://pdbp.ninds.nih.gov/), Biofind (https://biofind.loni.usc.edu/), and Parkinson’s Progression Markers Initiative (PPMI; https://www.ppmi-info.org/). In addition, we examined the association between *PTRHD1* and PD on a large European cohort of individual-level GWAS data including 14,671 PD cases and 17,667 controls from the International Parkinson’s Disease Genomics Consortium (IPDGC). Finally, we assessed summary statistics from the most recent GWAS meta-analyses on PD risk (Nalls et al., 2019) and age at onset (AAO) (Blauwendraat et al., 2020) excluding 23anMe data. In particular, the meta-analysis on PD risk consists of 15,056 PD cases, 18,618 UK Biobank proxy-cases (i.e., subjects with a first degree relative with PD) and 449,056 controls, whereas the meta-analysis on PD AAO includes 17,996 PD patients.

### 2.2 Genetic data

WGS data, processing and quality control pipelines can be found at www.amp-pd.org. Quality control for GWAS data was performed as previously described (Nalls et al., 2019). For both genotyping and sequencing datasets, *PTRHD1* region was annotated using ANNOVAR. To specifically analyze rare variants from WGS data, we selected variants with a minor allele frequency (MAF) <0.01. Allele frequency was calculated using PLINK v1.9 (Chang et al., 2015).

### 2.3 Statistical analysis

To test the association between variants in the *PTRHD1* region and PD, we performed Fisher exact test and logistic regression in WGS and imputed individual-level genotyping data adjusted for age, sex and principal components to account for population substructure, including variants within *PTRHD1* region +/- 100 Kb. Furthermore, summary statistics from the meta-analysis on PD AAO (Blauwendraat et al., 2020) and PD risk (Nalls et al., 2019) was investigated. Finally, as the *PTRHD1* variants analyzed in the reported family studies were all rare, we evaluated the enrichment of *PTRHD1* rare variants in WGS data using burden and kernel tests. The analyses on rare variants were first performed on all *PTRHD1* rare variants that passed quality control, and then repeated only on the nonsynonymous rare variants. Bonferroni correction for multiple comparisons was applied as needed. Association analyses were performed using PLINK v1.9 and burden analyses using software package RVTESTS v.2.1.0 (Zhan et al., 2016).

## 3. Results

In the WGS data we identified 1829 variants in the *PTRHD1* region +/-100 Kb, of which 29 were found within the *PTRHD1* gene. Among these, two were coding variants and 13 were identified as rare variants (MAF < 0.01). After Bonferroni correction, both logistic regression and Fisher test showed no association between *PTRHD1* variants and PD (Supplementary Tables S1-S2). We repeated the analyses using GWAS individual-level data and detected 511 variants in the *PTRHD1* region +/-100 Kb, with 14 variants in *PTRHD1* gene, none of which were coding. Similarly, the results showed no significant association between *PTRHD1* and PD (Supplementary Tables S3-S4). In addition, the analyses using the PD meta-analysis summary statistics confirmed such lack of association of *PTRHD1* with PD risk as well as with PD AAO (Figure 1). Finally, burden and kernel tests in WGS data did not show an enrichment of *PTRHD1* rare variants in PD, compared to controls (Supplementary Table S5). As the previously identified *PTRHD1* variants were all homozygous, we also assessed the number of homozygous/compound heterozygous carriers of at least one *PTRHD1* coding variant, showing similar proportions of such carriers between PD patients and controls (Supplementary Table S6). Of note, the three *PTRHD1* variants reported in the previous studies on Iranian and African families (Jaberi et al., 2016; Khodadadi et al., 2017; Kuipers et al., 2018) were not found in our dataset.

**Fig. 1.**
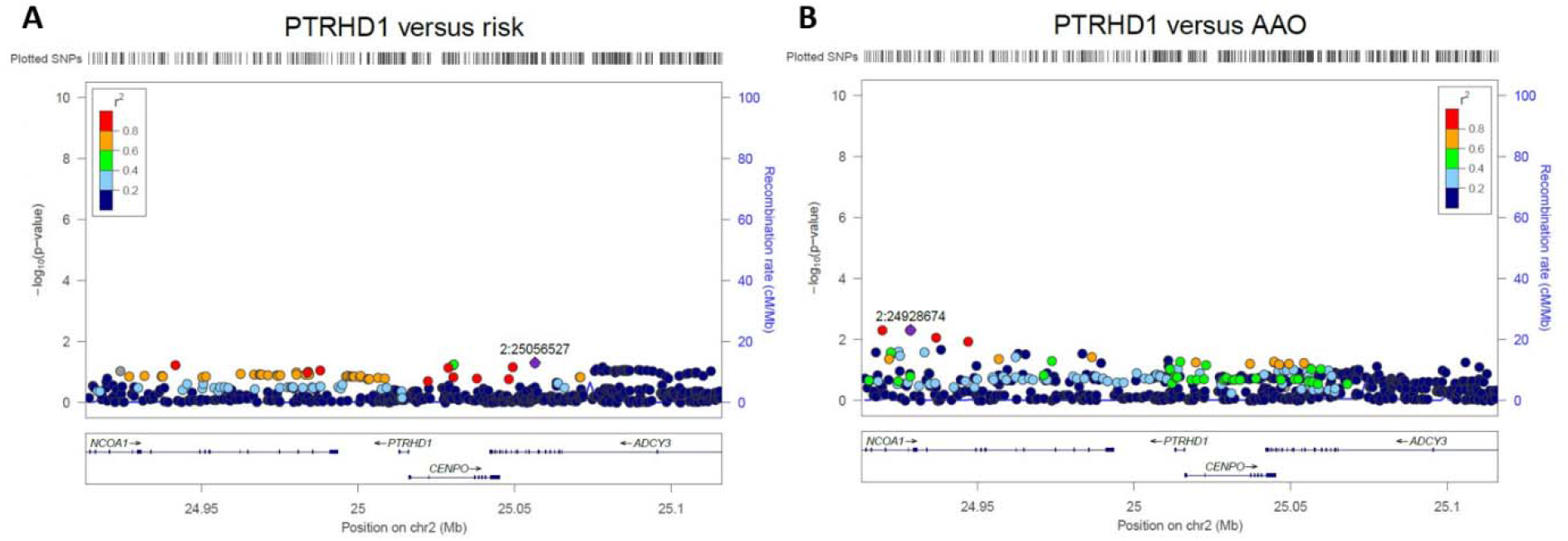
(A) Locus zoom plot of *PTRHD1* variants versus PD risk. (B) Locus zoom plot of *PTRHD1* variants versus PD AAO. The position of the variants on the chromosome 2 (x axis) is plotted against the log10-scaled p-values (left y axis). The most strongly associated SNP is indicated by a purple diamond. Pairwise linkage disequilibrium scores are defined by different colors, explained by the legend on the upper right corner. The right vertical axis indicates the regional recombination rate (cM/Mb). Abbreviations: chr, chromosome; cM, centimorgan; Mb, Megabase; PD, Parkinson’s disease; SNP, single nucleotide polymorphism.

## 4. Discussion

Herein, we did not find evidence for an association between *PTRHD1* and PD in the European population. There are several reasons that might explain the discrepancy between our results and the previous findings on *PTRHD1* in parkinsonian families (Jaberi et al., 2016; Khodadadi et al., 2017; Kuipers et al., 2018): 1) The findings from the family studies might not replicate on a large scale, suggesting that *PTRHD1* variants might be either extremely rare in PD or not associated at all; 2) *PTRHD1* carriers in the reported family studies manifested atypical parkinsonism with early-onset intellectual impairment, whereas our study was performed on patients with typical PD. Therefore, *PTRHD1* variants might be associated with rare forms of atypical parkinsonism, rather than typical PD; 3) The family studies were performed in Iranian and African subjects, whereas our study was performed in European patients. These ethnic differences might as well explain the discrepancy between our and the three previous family studies on *PTRHD1*. Our findings are further supported by a recent study conducted on PD families and early onset PD patients in a Taiwanese population (Chen et al., 2020). The study demonstrated an absence of *PTRHD1* pathogenic coding variants in PD patients.

In conclusion, our results do not provide evidence for an association between *PTRHD1* and PD. Larger studies on non-European ancestry populations and family trios are required to clarify the role of *PTRHD1* in PD and atypical parkinsonism.

## Data Availability

The code used in the current study is available on our GitHub page.
https://github.com/ipdgc/IPDGC-Trainees/blob/master/PTRHD1_IPDGC.md

## 5. Acknowledgements

We would like to thank all of the subjects who donated their time and biological samples to be a part of this study. We also would like to thank all members of the International Parkinson’s Disease Genomics Consortium (IPDGC). For a complete overview of members, acknowledgements and funding, please see http://pdgenetics.org/partners. This work was supported in part by the Intramural Research Programs of the National Institute of Neurological Disorders and Stroke (NINDS), the National Institute on Aging, and the National Institute of Environmental Health Sciences both part of the National Institutes of Health, Department of Health and Human Services; project numbers 1ZIA-NS003154, Z01-AG000949-02 and Z01-ES101986. In addition, this work was supported by the Department of Defense (award W81XWH-09-2-0128), and The Michael J Fox Foundation for Parkinson’s Research. Data used in the preparation of this article were obtained from the AMP PD Knowledge Platform. For up-to-date information on the study, visit https://www.amp-pd.org. AMP PD – a public-private partnership – is managed by the FNIH and funded by Celgene, GSK, the Michael J. Fox Foundation for Parkinson’s Research, the National Institute of Neurological Disorders and Stroke, Pfizer, and Verily. We would like to thank AMP-PD for the publicly available whole-genome sequencing data, including cohorts from the Fox Investigation for New Discovery of Biomarkers (BioFIND), the Parkinson’s Progression Markers Initiative (PPMI), and the Parkinson’s Disease Biomarkers Program (PDBP). This work utilized the computational resources of the NIH HPC Biowulf cluster (http://hpc.nih.gov). ZGO is supported by the Fonds de recherche du Québec - Santé (FRQS) Chercheurs-boursiers award, in collaboration with Parkinson Quebec, and by the Young Investigator Award by Parkinson Canada.

